# Association of Assisted Reproductive Technologies with offspring cord blood DNA methylation across cohorts

**DOI:** 10.1101/2020.06.18.20134940

**Authors:** Doretta Caramaschi, James Jungius, Christian M. Page, Boris Novakovic, Richard Saffery, Jane Halliday, Sharon Lewis, Maria C. Magnus, Stephanie J. London, Siri E. Håberg, Caroline L. Relton, Deborah A. Lawlor, Hannah R. Elliott

## Abstract

**Study question:** Is DNA methylation at birth associated with having been conceived by assisted reproductive technologies (ART)?

**Summary answer:** This study shows does not provide strong evidence of an association of conception by ART with variation in infant blood cell DNA methylation.

**What is known already:** Assisted reproductive technologies (ART) are procedures used to help infertile/subfertile couples conceive. Due to its importance in gene regulation during early development programming, DNA methylation and its perturbations associated with ART could reveal new insights into the biological effects of ART and potential adverse offspring outcomes.

**Study design:** We investigated the association of DNA methylation and ART using a case-control study design (N=205 ART cases and N=2439 non-ART controls in discovery cohorts; N=149 ART cases and N=58 non-ART controls in replication cohort).

**Participants/materials, settings, method:** We assessed the association between ART and DNA methylation at birth in cord blood (205 ART conceptions and 2439 naturally conceived controls) at >450000 CpG sites across the genome in two sub-samples of the UK Avon Longitudinal Study of Parents and Children (ALSPAC) and two sub-samples of the Norwegian Mother, Father and Child Cohort Study (MoBa) by meta-analysis. We explored replication of findings in the Australian Clinical review of the Health of adults conceived following Assisted Reproductive Technologies (CHART) study (N=149 ART conceptions and N=58 controls).

**Main results and the role of chance:** The ALSPAC and MoBa meta-analysis revealed evidence of association between conception by ART and DNA methylation (false-discovery-rate-corrected p-value < 0.05) at 5 CpG sites which are annotated to 2 genes. Methylation at 3 of these sites has been previously linked to cancer, aging, HIV infection and neurological diseases. None of these associations replicated in the CHART cohort. There was evidence of a functional role of ART-induced hypermethylation at CpG sites located within regulatory regions as shown by putative transcription factor binding and chromatin remodelling.

**Limitations, reasons for cautions:** While insufficient power is likely, heterogeneity in types of ART and between populations may also contribute. Larger studies might identify replicable variation in DNA methylation at birth due to ART.

**Wider implications of the findings:** ART-conceived newborns present with divergent DNA methylation in cord blood white cells. If these associations are true and causal, they might have long-term consequences for offspring health.

## Introduction

The number of children conceived using assisted reproductive technologies (ART) has increased steadily over the last four decades. ART are defined here as including procedures such as *in vitro* fertilisation (IVF), including its modified versions such as intra-cytoplasmatic sperm injection (ICSI) or gamete intrafallopian transfer, where there is manual handling of gametes and exposure to the artificial *in vitro* and hormone-stimulated *in vivo* environments, as well as medically assisted reproduction procedures (Zegers-Hochschild et al. 2017), which might also use hormones to stimulate oocyte production but do not usually handle gametes. ART may subject gametes and early embryos to environmental stress, which may impact the development of the fetus, as shown for instance by disruptions in the expression of imprinted genes (Sakian et al. 2015). The timing of epigenetic reprogramming, coinciding with ART interventions and the susceptibility of DNA methylation to environmental stresses, has led to suggestions that ART interventions may alter the offspring’s DNA methylation (Canovas et al. 2017).

Earlier studies investigating the association of ART and DNA methylation at birth have focused on imprinted genes and found mixed results which point to weak evidence of any association when meta-analysed together (Lazaraviciute et al. 2014). Other studies have included other non-imprinted candidate genes and found some associations in cord and placental cells (Katari et al. 2009; Tierling et al. 2010; Song et al. 2015). More recent genome-wide studies found differences in neonatal blood cells between ART and controls (Melamed et al. 2015; Estill et al. 2016; Castillo-Fernandez et al. 2017; El Hajj et al. 2017; Novakovic et al. 2019) although other similar studies have found little evidence (Gentilini et al. 2018; Choufani et al. 2019), perhaps owing to small sample sizes, comparisons with different ART treatments across some studies and consequently insufficient power to detect true associations. Only one study have included more than 100 ART conceptions (Novakovic et al. 2019), and, although it found several associations, it did not control for potential confounding from maternal characteristics.

Whilst different ARTs have been associated with a wide range of adverse perinatal and longer-term offspring outcomes (Kallen et al. 2010; Hansen et al. 2013; Chen and Heilbronn 2017; Elias et al. 2020), a large recent study with follow-up to 22-35 years, did not confirm the associations with longer-term outcomes associations (Halliday et al. 2019). Moreover, the extent to which any associations with adverse outcomes are related to multiple pregnancies, the effect of underlying causes of infertility, selection bias or confounding is unclear. In relation to adverse perinatal outcomes such as pregnancy loss, preterm birth and small for gestational age, increasing evidence suggests that multiple pregnancies explain much of the increased risk (Nelson and Lawlor 2011; Lawlor and Nelson 2012). Recent evidence from the European Surveillance of Congenital Anomalies (EUROCAT) suggests that a substantial proportion of associations with congenital anomalies may be due to underlying causes of infertility rather than ART *per se* (Davies et al. 2012; Levi Setti et al. 2016). However, these and studies of ART with longer-term offspring outcomes, often rely on large-scale clinical and administrative datasets with minimal data on potential confounders. Studies that are based around ART cohorts usually have more detailed information on potential confounders but may suffer from selection bias because of selecting spontaneous conception controls from volunteers (often with low response rates), or friends and family members of the couple undergoing ART, who are more like the ART couples than the general population from which those couples came (Halliday et al. 2014). Exploring the association of ART with infant epigenetic variation in birth cohorts that did not select any participants on the basis of how infants were conceived could contribute to understanding possible causal effects of ART on offspring health. First, we would expect less selection bias as both the ART and comparison group were selected into the study in the same way. Second, these birth cohorts will have detailed information on a range of potential confounders. The identification of associations of ART with differential DNA methylation in genes that have established associations with health outcomes could be used to prioritise offspring disease outcomes for more detailed causal analyses that might be mediated by DNA methylation. Interrogating DNA methylation and its potental role as a mediating mechanism linking ART to health consequeces in the offspring may provid insights into the biological pathways involved.

The aim of this study was to determine the association between having been conceived through ART and genome-wide variation in cord blood DNA methylation at >450000 CpG sites using the Illumina Methylation 450K BeadChip across multiple cohorts. Building on previous studies, we have used a multi-cohort approach with meta-analysis and replication, while interrogating the whole genome. We have also used functional analysis methods to attempt to characterise the biological implications of the findings in the context of long-term health outcomes.

## Material and Methods

The overall approach to our methods was to pool results from EWAS in two general population birth cohorts, the Avon Longitudinal Study of Parents and Children (ALSPAC) and the Norwegian Mother, Father, and Child Birth Cohort (MoBa), as discovery analyses. We then explored replication of any discovery associations in an independent study, the “Clinical review of the Health of adults conceived following Assisted Reproductive Technologies” (CHART) cohort.

### Study populations

#### Avon Longitudinal Study of Parents and Children (ALSPAC)

ALSPAC is a population birth cohort that enrolled 14,541 pregnancies from residents in and around the city of Bristol in the South West of the UK with expected delivery dates between 1991 and 1992 (Boyd et al. 2013; Fraser et al. 2013). The study website contains details of all the data that is available through a fully searchable data dictionary and variable search tool: (http://www.bristol.ac.uk/alspac/researchers/our-data/).

In ALSPAC, a subset of 1018 mother-offspring pairs had DNA methylation measured within the Accessible Resource for Integrated Epigenomics Studies (ARIES) (http://www.ariesepigenomics.org.uk/) (Relton et al. 2015). ALSPAC-ARIES participants were selected based on the availability of DNA samples at two time points for mothers and three time points for the offspring (Relton et al. 2015). After quality control checks and exclusion of multiple pregnancies N=857 pairs had reliable cord blood DNA methylation data to include in this study. After removing N=21 participants without ART data and further N=52 participants without all the covariates, we had a sample of N=784 (77% of the original ARIES samples). In comparison to the overall ALSPAC cohort the subset of ARIES mother-offspring pairs are slightly older, more likely to have a non-manual occupation and less likely to have smoked during pregnancy (Relton et al. 2015).

A second ALSPAC subset, the ALSPAC-ART subsample, also had cord blood DNA methylation measured. This subset consisted of additional ALSPAC participants (ART and non-ART controls) selected to maximise the number of children conceived through ART and hence increase power. 178 ART-conceived infants with cord blood samples and 241 non-ART controls were selected for epigenome wide DNA methylation analyses. The 241 non-ART conceived participants were not a random sample but consisted of participants selected for other epigenetic experiments that were run in the lab at the same time as the ART cases), based on other phenotypes (arthritis, diabetes, eating disorder, pre-eclampsia, hypertension, body mass index (BMI), child autism and child undescended testicle). The distribution of these traits across the ART and non-ART participants is shown in Supplementary Table 1. After removing those without all the covariates, there were N=345 participants (82% of the additional subset; N=155 ART and N=190 non-ART). Written informed consent was obtained from all participants. The protocols and types of chip used for measurement of epigenome wide DNA methylation were the same for both ALSPAC samples. Ethical approval for the ALSPAC data was granted by the ALSPAC law and ethics committee and local Research Ethics Committees. The details of the ethical approval at each stage of the study can be found in the ALSPAC website (http://www.bristol.ac.uk/alspac/researchers/research-ethics/).

#### The Norwegian Mother and Child Cohort Study (MoBa)

Participants represent two subsets of mother-offspring pairs from the national Norwegian Mother and Child Cohort Study (MoBa), a prospective population-based pregnancy cohort study conducted by the Norwegian Institute of Public Health (Magnus et al. 2006; Magnus et al. 2016). The years of birth for MoBa participants ranged from 1999-2009 and the cohort includes more than 114000 children, 95000 mothers and 75000 fathers. Two subsets of children also have cord blood DNA methylation available. The two subsets are referred to here as MoBa1 and MoBa2. MoBa1 is a subset of a larger study within MoBa that includes a cohort random sample and newborns who developed asthma by age three years (Haberg et al. 2011), of which N=1068 children had cord blood DNA methylation available after quality control exclusions (Joubert et al. 2012). MoBa2 consists of a further 685 participants who were also a random cohort sample and newborns who developed asthma that was still present at age seven (157 with asthma) (Reese et al. 2019). MoBa2 participants did not overlap with those in MoBa1 and the protocol and type of chips used for DNA methylation analyses were the same in both subsets. Cord blood DNA methylation data after quality control exclusions were available in N= 680 MoBa2 children. MoBa1 and MoBa2 data were pooled together and N=1518 (22 conceived by ART) of these had data on ART and all the covariates included in the analysis.

Ethical approval for MoBa data was granted by Regional Committee for Ethics in Medical Research of South/East Norway, the Norwegian Data Inspectorate, and the Institutional Review Board of the National Institute of Environmental Health Sciences. Written informed consent was obtained from all MoBa participants.

#### ART Assessment

ALSPAC participants were sent a postal questionnaire at 12 weeks gestation that asked a two-part question: i) “Did you use any treatments to help conceive this pregnancy?”, a binary yes/no response, and ii) “If yes which one?”, a text answer. We used the first question to create a binary ART variable for the main analysis. We used the text response to the second question to perform a sensitivity analysis where the ART category included only mothers who conceived using IVF, intrauterine insemination, ovulation induction by hormones or male infertility treatments, whilst excluding those who reported using natural monitoring, other fertility treatments, other treatment (not fertility), no treatment or unknown and those with mismatches between the two questions (N=7 removed in ALSPAC-ARIES and N=28 removed in ALSPAC-ART).

In MoBa ART information was obtained from the medical birth registry. The notification form completed by the midwife attending the delivery indicated whether the pregnancy was conceived by IVF, ICSI or other/unknown ART methods, as a single response.

#### DNA methylation

In the ALSPAC-ARIES and ALSPAC-ART subsamples, DNA was extracted from previously collected and frozen cord blood. DNA was then bisulphite converted using Zymo EZ DNA MethylationTM kit (Zymo, Irvine, CA) and methylation was measured at 485,512 CpG sites using the Illumina Infinium HumanMethylation450K BeadChip assay according to manufacturers’ protocols (Relton et al. 2015). The methylation arrays were scanned with an Illumina iScan and the initial quality was checked with GenomeStudio (version 2011.1). The methylation data pre-processed using *meffil* package (Min et al. 2018). The process included quantile normalisation and quality checks such as sex chromosomes mismatches and genotype mismatches.

In MoBa1 umbilical cord blood samples were collected and frozen at birth at −80°C. All biological material was obtained from the Biobank of the MoBa study (Paltiel et al. 2014). Bisulfite conversion was performed using the EZ-96 DNA Methylation kit (Zymo Research Corporation, Irvine, CA) and DNA methylation was measured at 485,577 CpGs in cord blood using the Illumina Infinium HumanMethylation450 BeadChip array (Joubert et al. 2012). Raw intensity (.idat) files were handled in R using the *minfi* package to calculate the methylation level at each CpG as the beta-value and the data was exported for quality control and processing. Similar sample specific quality control was performed in MoBa1 and MoBa2, as follows: control probes (N=65) and probes on X (N=11 230) and Y (N=416) chromosomes were excluded in both datasets. Remaining CpGs missing > 10% of methylation data were also removed (N=20 in MoBa1, none in MoBa2). Samples indicated by Illumina to have failed or have an average detection p-value across all probes < 0.05 (N=49 MoBa1, N=35 MoBa2) and samples with gender mismatch (N=13 MoBa1, N=8 MoBa2) and GWAS outliers (N = 6 MoBa1, N = 4 MoBa2) were also removed. For MoBa1 and MoBa2, we accounted for the two different probe designs by applying the intra-array normalization strategy Beta Mixture Quantile dilation (BMIQ) (Bibikova et al. 2011). After quality control exclusions, the sample sizes of the cord blood DNA methylation data (including with missing ART and covariates) were 1056 for MoBa1 and 680 for MoBa2. To account for differences between MoBa1 and MoBa2, we also adjusted for the different design variables, such as sample pull or selection factors (MoBa2) with a fixed effect variable.

#### Epigenome-wide association study (EWAS) meta-analysis

We estimated the association between ART and cord blood DNA methylation at all the CpGs that passed QC in ALSPAC-ARIES, ALSPAC-ART and MoBa by running linear regression models of ART (exposure) on DNA methylation (outcome) using the *CpGassoc* package in R (Barfield et al. 2012). To control for batch, we included a term for bisulphite treatment plate. We used an algorithm and a reference panel that is suitable for cord blood (Bakulski et al. 2016) to calculate cell type composition and added estimated cell counts as covariates. Additionally, we adjusted for infant sex at birth and potential confounding by maternal age at delivery, maternal BMI and maternal smoking during the first trimester of pregnancy. In ALSPAC, maternal pre-pregnancy weight, height and smoking were obtained through self-report in questionnaires completed in the first trimester of pregnancy whereas maternal age at delivery and infant sex were extracted from obstetric records. Smoking during the first trimester was categorised into a binary variable of “Did not smoke” and “Smoked”. In MoBa, for both datasets, information maternal pre-pregnancy weight height and smoking was obtained through self-report questionnaires completed at around 17 weeks of gestation; maternal age at delivery and infant sex were obtained from the birth registry records. Smoking was categorised in a three-level variable, “non-smoker”, “stopped early in pregnancy”, and “smoked throughout pregnancy”.

A meta-analysis of the summary statistics of the EWAS results from each study sample (ALSPAC-ARIES, ALSPAC-ART and MoBa) was carried out using METAL (version 2011-03-25) (Willer et al. 2010), using a fixed effects model with inverse variance weighting (Rice et al. 2018). Since in the MoBa EWAS the X and Y chromosome were omitted, the meta-analysis excluded CpGs on these chromosomes.

#### Replication in an independent cohort

We sought replication of results for all CpG sites with p_FDR_ <0.05 in the meta-analysis EWAS in an independent study sample, the “Clinical review of the Health of adults conceived following Assisted Reproductive Technologies” (CHART) cohort. CHART is an Australian cohort of N=547 adults conceived by IVF and N=549 naturally conceived controls (Lewis et al. 2017). In a subsample of N=193 ART-conceived and N=86 non-ART adults who gave their consent for epigenetic analyses, DNA methylation was measured in DNA isolated from neonatal blood spots (Guthrie spots) that were collected at 2-3 days after birth using the Illumina InfiniumMethylationEPIC BeadChips array. Pre-processing was carried out using *MissMethyl* and *minfi* R packages and normalization was performed using SWAN (Maksimovic et al. 2012). Quality checks involved the exclusion of samples with mean detection p-value>0.01, probes with detection p-value>0.01, those associated with SNPs and cross-reactive probes. Cell composition was estimated using the Bakulski cord blood cell reference method (Bakulski et al. 2016). Linear regression modelling was performed using *limma* R package and included sample plate, cell proportions (six cell types), infant sex and maternal age at delivery covariates (data were not available in this study to adjust for potential confounding by maternal pre-pregnancy BMI or pregnancy smoking). After quality control exclusions there were 146 ART cases and 58 controls with data on ART and covariates to use in this analysis. Ethical approval for the CHART study was granted by the Royal Children’s Hospital Human Research Ethics Committee (RCH HREC Project 33163). All participants for this study gave written informed consent.

#### Biological characterisation

CpG sites associated with ART were compared to those listed in the EWAS catalog (http://www.ewascatalog.org/) in July 2019. Studies indexed by the EWAS catalog included at least 100,000 CpG sites in analyses and had sample sizes of at least 100 individuals.

Gene ontology enrichment (Ashburner et al. 2000; The Gene Ontology 2017) for the first 100 CpG sites from the meta-EWAS ranked by ascending p-value was conducted using the R package *missMethyl* (Phipson et al. 2016). This method takes into account the differing number of probes per gene found on the array which can otherwise bias results (Geeleher et al. 2013). Since hyper- and hypo-methylation at CpG sites in our analysis were likely to be biologically distinct we stratified our enrichment analysis accordingly.

Enrichment analyses were conducted using the R package LOLA (Sheffield and Bock 2016). Input was genomic coordinates of the first 100 CpG sites from the meta-EWAS ranked by ascending p-value and the background set was the genomic coordinates all array CpG sites included in the meta-EWAS. The LOLA core region set was used to test for enrichments. As above, sites were stratified into hyper- and hypo-methylated CpG sites. For each LOLA analysis, we filtered results retaining enriched region sets only when the support (i.e. number of regions overlapping) >=5 and the q-value was <0.05.

To further identify enrichment of ART-associated CpG sites at cell type specific histone modifications and DNaseI hypersensitivity sites (DHS) we used eFORGEv2.0 (Breeze et al. 2016). H3 marks and DHS were analysed from the consolidated Roadmap Epigenomics data set. Input was the first 100 CpG sites from the EWAS when ranked by ascending p-value. The background set of CpG sites was all CpG sites on the array analysed in the meta-EWAS. As above, sites were stratified into hyper- and hypo-methylated CpG sites.

The genes mapped to the CpGs associated with ART at P_FDR_<0.05 and in DMRs were searched for published associations in the GWAS Catalog (https://www.ebi.ac.uk/gwas/, v1.0.2 date accessed: 04/06/2020) (Buniello et al. 2019).

#### mQTLs analysis

For each CpG associated with ART in the single-site EWAS at P_FDR_<0.05, we searched for mQTLs in the mQTL database (www.mqtldb.org, date accessed: 10/06/2020) (Gaunt et al. 2016).

## Results

### Study sample characteristics

The characteristics of the study participants for ALSPAC-ARIES, ALSPAC-ART and MoBa are shown in Table 1 and Supplementary Table 2. ALSPAC-ART and MoBa included a higher proportion of smokers during the first trimester and female offspring compared to ALSPAC-ARIES. Mean birthweight, gestational length and maternal pre-pregnancy BMI were similar across the three study samples. In all three samples mothers of infants conceived via ART were older and in ALSPAC-ART and MoBa ART-conceived infants were slightly more likely to be female. In ALSPAC-ART, and to a weaker extent ALSPAC-ARIES, mothers of ART-conceived infants were less likely to smoke and had lower BMI, which may reflects UK guidelines that restrict some ART procedures to women with a healthy BMI and who do not smoke at the time of treatment.

**Table 1.** Characteristics of the three study samples used in the EWAS meta-analysis.

|  | ALSPAC-ARIES<br>N=784 | ALSPAC-ART<br>N=345 | MoBa<br>N=1518 |
| --- | --- | --- | --- |
| ART (%) | 28 (3.6) | 155 (44.9) | 22 (1.5) |
| Smoked first trimester (%) | 100 (12.8) | 83 (24.1) | 415 (27.3) |
| Offspring gender at birth (% Female) | 398 (50.6) | 143 (41.4) | 706 (46.5) |
| Maternal age (yrs)<br>(Mean (SD)) | 29.65 (4.41) | 28.84 (4.83) | 29.96 (4.31) |
| Maternal BMI (kg/m<sup>2</sup>) (Mean (SD)) | 22.92 (3.80) | 24.52 (6.57) | 24.12 (4.32) |
| Birthweight (g) (Mean (SD)) | 3419 (481) | 3364 (606) | 3648 (541) |
| Gestational age (wks) (Mean (SD)) | 39.70 (1.48) | 39.33 (1.86) | 39.48 (1.61) |

### EWAS meta-analysis

Figure 1 and Table 2 summarise the results of the epigenome-wide association study meta-analysis adjusted for gender at birth, maternal age at delivery, maternal BMI and maternal smoking during pregnancy. There were 5 CpG sites associated with ART at the genome-wide level (P_FDR_<0.05), 4 of which surpassed the threshold for Bonferroni corrected p-values (P_Bonferroni_<0.05). These were hyper-methylated sites. The effect sizes were lower than 1% absolute difference in methylation between the two groups. The direction of effect was concordant across studies for 3 of the 5 sites and it was concordant for 4 of the 5 sites between the two ALSPAC subsamples. Three of the CpG sites were located in known genes, including two sites that were annotated to the same gene coding for arrestin domain-containing protein 1 (*ARRDC4*). In all three study samples methylation at *ARRDC4* was <1% higher in ART cases compared to non-ART controls at unadjusted p-value<0.05.

**Figure 1.**
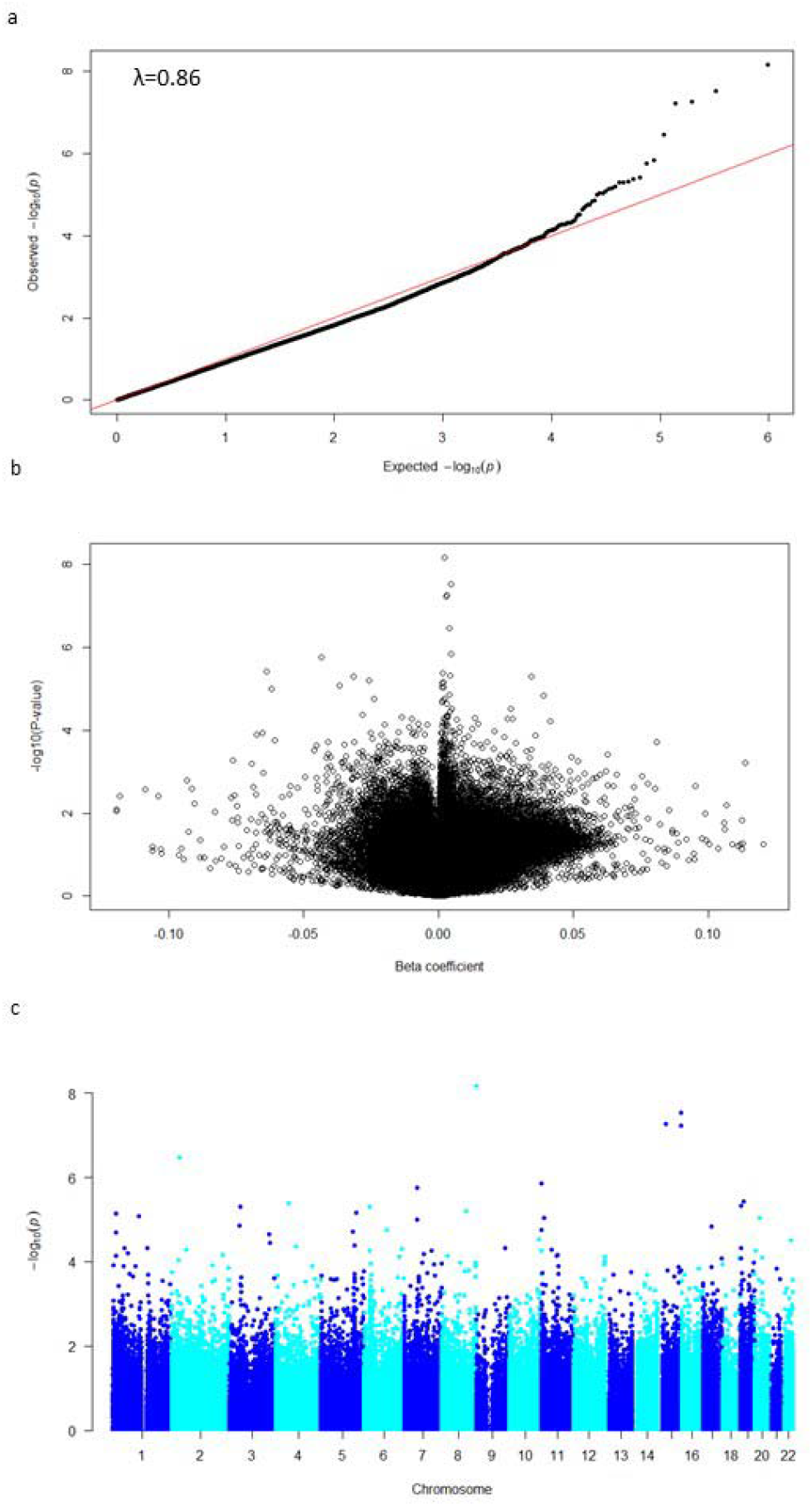
Association between cord blood DNA methylation and ART estimated by the meta-analysis. (a) Quantile-quantile plot of the observed vs expected distribution of p-values; (b) Volcano plot of the p-values vs the beta coefficients. λ= genomic inflation factor calculated using the median method. (c) Manhattan plot of the distribution of p-values across chromosomes.

**Table 2.**
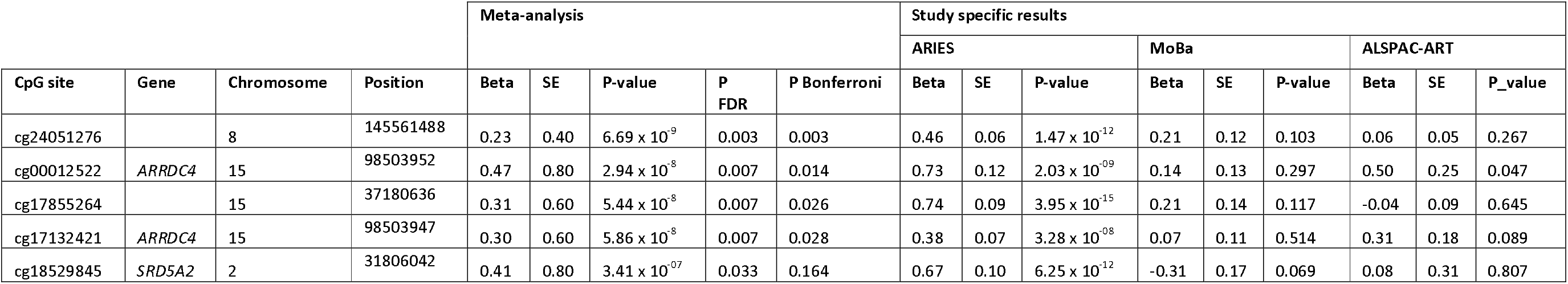
Results of the epigenome-wide association study meta-analysis (CpG sites at P_FDR_<0.05). Beta regression coefficients are reported as percent change in methylation.

EWAS analyses conducted in ALSPAC-ARIES and ALSPAC-ART with restriction to participants who reported specific forms of ART (IVF, intrauterine insemination, ovulation induction by hormones or male infertility treatments) and adjusted for the same covariates as the main analysis showed similar results to the main analyses with high correlation between the regression beta coefficients (R^2^= 0.86 for ALSPAC-ARIES and R^2^=0.95 for ALSPAC-ART).

### Replication in CHART

In the CHART sample, there was no clear evidence of association at any of the 5 CpG sites identified in the discovery meta-analysis (Supplementary Table 3).

### Biological characterisation

Three of 5 sites associated with ART at genome-wide level in our discovery sample, were previously reported to be associated with other phenotypes according to the EWAS Catalog (See supplementary Table 4). cg18529845 and cg17855264 were associated in multiple EWAS studies of cancer, ageing and HIV infection. cg24051276 was associated with rheumatoid arthritis, dementia and palsy.

No GO terms were shown to be enriched after accounting for multiple testing. In the hypermethylated probe set the most highly enriched terms were biological processes linked to modulation by host of viral process and androgen metabolism (See Supplementary Table 5).

To identify enrichment of ART-associated sites within gene and gene regulatory regions we utilised eFORGE and LOLA. Using LOLA, enrichment was identified amongst the set of CpG sites that were hypermethylated in ART cases compared to controls (n=69 of 100 sites included). Hypermethylated CpGs were enriched for CpG islands (q= 0.0011) (see Supplementary Table 6). They were also enriched for EZH2 transcription factor binding sites based on ENCODE data. This transcription factor is associated with methylation of H3K27 chromatin leading to chromatin compaction (Laugesen et al. 2019). LOLA analysis also showed that hypermethylated CpG sites were strongly enriched amongst H3K4me2, H3K4me3, H3K9me3 and H3K4me1 in breast and prostate tissues using data derived from the cistrome project.

eFORGE was used to identify cell type specific DHS in tissues profiled by the NIH Roadmap Epigenomics consortium (see Supplementary Table 7). Enrichment was observed for the hypermethylated set. The strongest enrichment identified for DHS was found in embryonic stem cells. The histone modifications H3K9me3 and H3K9me1 were strongly enriched in multiple tissues, including embryonic stem cells, blood, ovary, stem and fetal cells. H3K9me3 is associated with decreased accessibility of chromatin and is pivotal in lineage commitment during organogenesis and for maintaining cell lineage in differentiated cells (Nicetto et al. 2019). H3K4 methylation was also identified using LOLA and is typically detected in promoter regions of active genes (Barski et al. 2007).

The ‘GWAS Catalog’ search showed that a genetic variant in *SRD5A2*, whose methylation was associated with ART, had previously been found to be associated with male baldness (Supplementary Table 8).

### mQTL analysis

To explore the potential health consequences of ART, we searched for *cis*-mQTLs of the top 5 CpGs from the meta-analysis by looking at SNPs associated with the CpGs in the mQTLdb database (www.mqtldb.org). We were not able to identify any suitable mQTL from this analysis as all the SNPs associated with the 5 top CpGs were located in *trans* (>1 Kb from the CpG site).

## Discussion

We conducted an epigenome-wide association study meta-analysis of the association of ART with cord blood DNA methylation across two European birth cohorts, ALSPAC and MoBa. The results showed evidence of association between ART and DNA methylation at 5 CpG sites. By interrogating online databases, we also found evidence that some of the methylation differences associated with ART were associated with diseases and health-related traits in adulthood. At the molecular level, ART-linked hypermethylation occurred in regulatory sites involved in chromatin remodelling. However, we could not confirm whether ART associations with differential methylation at these 5 CpGs were causal (or might be explained by confounding, including by the underlying pathology resulting in infertility). Nor could we establish whether ART-associated methylation changes were causally linked to diseases and health-related traits using Mendelian randomization.

Several previous studies have explored differential DNA methylation in infant blood cells related to conception by ART using either candidate or EWAS approaches. Most of these have had smaller sample sizes than our study and/or have not sought replication in an independent study. In a recent study conducted on the CHART cohort used as replication in our study (Novakovic et al. 2019), 18 CpG sites were associated with ART, 9 of which were available in our study. In our meta-analysis, at 4 of these 9 CpGs the direction of the effects was concordant (including at *CHRNE*) and with low p-values (0.07 to 0.006) (Supplementary Table 9), thus providing some evidence of consistency across studies, although none of these 9 CpGs had p-values<0.006 (p=0.05 Bonferroni-corrected to take into account 9 multiple comparisons). Differences between our study and this one may be explained by differences in the adjustment for confounding factors (we adjusted for batch, cell-type, maternal pre-pregnancy BMI, pregnancy smoking, age at delivery and infant sex, whereas the published study adjusted for batch, cell-type and infant sex), the different sample used (blood in our study and Guthrie spots in the published study) and/or differences in ART definition (self-reported in our study and from hospital records in the published study). However, given that the two studies were broadly similar in population, methods and size the lack of consistency between the two may be because the findings in both studies are due to chance and do not reflect real replicable associations. This would be consistent with the lack of replication of our findings in the CHART study.

Most of the CpGs identified in this study are located in the proximity of known genes. We found evidence of hypermethylation at the transcription start site of the *SRD5A2* gene, coding for steroid 5-alpha reductase 2, an enzyme involved in androgen metabolism and male sexual development. Since our study is observational, we cannot rule out that an impairment in this gene at the parental level was present before having received ART procedures and rather than its consequence and could be related to parental sub-fertility and not ART treatment *per se*. The lack of this and the other sites that were differentially methylated in our discovery sample may be due to the different sample collections (cord blood in our discovery meta-analysis and Guthrie dry blood spots collected at 2-4 days in CHART). They could also be due to different outcome definitions. In our European meta-analyses exposure to any ART was reported by the mother and the question in ALSPAC was quite vague, though sensitivity analyses restricting to those who described an ART procedure were similar to our main analyses, whereas in CHART only those conceived by IVF were included in the ART group and this was obtained from hospital records. However, the most likely interpretation is that these additional associations are driven by chance.

Our study has several strengths compared to other previously published studies. By combining participants from different studies, we have achieved a larger sample size. We also adjusted for potential confounding maternal characteristics (maternal pre-pregnancy BMI, pregnancy smoking and age at delivery), whereas most previous studies had either smaller sample sizes or did adjust for fewer confounders. We conducted extensive biological characterisation using freely available databases and an mQTL look-up to attempt to determine potential causal links to disease and health-related traits. Finally, we explored replication in an independent sample.

This study has also some limitations. Firstly, due to the observational nature of the study it has not been possible to establish causality between ART conception and DNA methylation. Even though we adjusted for potential confounders the possibility of residual confounding, for example by the underlying cause of parental sub-fertility, remains. Furthermore, due to an insufficient number of mQTLs, our study did not provide the opportunity to conduct Mendelian randomization to explore causal effects between ART-associated methylation differences and disease risk. A more detailed mQTL catalog, and therefore source of genetic instrumental variables, might help to rectify this limitation in the future. In our discovery meta-analysis our main exposure was any ART and we were unable to explore whether different types of ART-associated differently with DNA methylation. In ALSPAC, participants were asked a very general question about having used any treatment to help them conceive and then had the opportunity to provide additional information in a text response, with some women suggesting their treatment was not ART and others not responding. However, results were very similar when we repeated analyses only focusing on those who reported a definite ART. In MoBa, data was obtained from the birth register, but the report was a single entry completed by a midwife that asked whether the conception was by ART (defined as IVF, ICSI or other ART) so we were unable to explore different types of ART.

Overall, we show that newborns conceived by ART procedures present with altered DNA methylation in cord blood white cells. However, we cannot distinguish whether these ART-associated methylation profiles are due to ART, reflect the underlying causes of subfertility, or are chance associations. If any are causal, we do not know whether there are long-term consequences for offspring health. As this is one of the largest studies to date, has adjusted for confounders and tried to explore replication, it highlights the need for larger studies and/or attempts to harmonise data across all studies and undertake a larger collaborative effort. There is also a need for studies with more detailed information on the type of ART, and on the cause of parental sub-fertility. However, to the best of our knowledge studies with such detailed exposure information and DNA methylation in large numbers are currently lacking. Additional follow-up studies with DNA methylation at older ages would also be useful to investigate a potential resolution of ART-induced DNA methylation variation over time as indicated by the CHART study (Novakovic et al. 2019). Current efforts in generating more mQTL databases such as the GoDMC consortium (http://www.godmc.org.uk/) might provide more mQTL for use in Mendelian randomisation to establish causal links between ART-related differential DNA methylation and future disease risks.

## Data Availability

The data underlying this article cannot be shared publicly for the privacy of individuals that participated in the study. The data will be shared on reasonable request to the individual cohorts. For ALSPAC, data are available according to the procedures listed at http://www.bristol.ac.uk/alspac/researchers/access/. For MoBa, data can be requested at https://www.fhi.no/en/studies/moba/for-forskere-artikler/research-and-data-access/.

## Author’s Contributions

D.C., J.J., H.R.E. data analysis, interpretation of results and manuscript preparation; C.M.P., B.N., data analysis and draft revisions; R.S., J.H. and S.J.L. data acquisition and draft revision; D.A.L. and C.L.R. conception/design of the work, data acquisition, interpretation of results and draft revisions. All authors provided critical input on the manuscript.

## Acknowledgements and Funding

The UK Medical Research Council and Wellcome (Grant ref: 102215/2/13/2) and the University of Bristol provide core support for ALSPAC. This study has also been supported by the US National Institute of Health (R01 DK10324), the European Research Council under the European Union’s Seventh Framework Programme (FP7/2007-2013) / ERC grant agreement no 669545, European Union’s Horizon 2020 research and innovation programme under grant agreement No 733206 (LifeCycle) and the NIHR Biomedical Centre at the University Hospitals Bristol NHS Foundation Trust and the University of Bristol. A comprehensive list of grants funding is available on the ALSPAC website (http://www.bristol.ac.uk/alspac/external/documents/grant-acknowledgements.pdf). Methylation data in the ALSPAC cohort were generated as part of the UK BBSRC funded (BB/I025751/1 and BB/I025263/1) Accessible Resource for Integrated Epigenomic Studies (ARIES, http://www.ariesepigenomics.org.uk). D.C., J.J., C.R. and H.R.E. are funded by the UK Medical Research Council (grant numbers: MC_UU_12013/1-2, MC_UU_00011/1, MC_UU_00011/5). B.N. is supported by an NHMRC (Australia) Investigator Grant (1173314). GWAS data was generated by Sample Logistics and Genotyping Facilities at Wellcome Sanger Institute and LabCorp (Laboratory Corporation of America) using support from 23andMe. We are extremely grateful to all the families who took part in this study, the midwives for their help in recruiting them, and the whole ALSPAC team, which includes interviewers, computer and laboratory technicians, clerical workers, research scientists, volunteers, managers, receptionists and nurses. This publication is the work of the authors and Doretta Caramaschi will serve as guarantor for the contents of this paper. The Norwegian Mother, Father and Child Cohort Study is supported by the Norwegian Ministry of Health and Care Services and the Ministry of Education and Research, NIH/NIEHS (contract no N01-ES-75558), NIH/NINDS (grant no.1 UO1 NS 047537-01 and grant no.2 UO1 NS 047537-06A1). For this work, MoBa 1 and 2 were supported by the Intramural Research Program of the NIH, National Institute of Environmental Health Sciences (Z01-ES-49019) and the Norwegian Research Council/BIOBANK (grant no 221097). This work was partly supported by the Research Council of Norway through its Centres of Excellence funding scheme, project number 262700. We are very grateful to all the participating families in Norway who take part in this on-going cohort study.

